# Reduced BNT162b2 mRNA vaccine response in SARS-CoV-2-naive nursing home residents

**DOI:** 10.1101/2021.03.19.21253920

**Authors:** David H. Canaday, Lenore Carias, Oladayo A. Oyebanji, Debbie Keresztesy, Dennis Wilk, Michael Payne, Htin Aung, Kerri St. Denis, Evan C. Lam, Brigid Wilson, Christopher F. Rowley, Sarah D. Berry, Cheryl M. Cameron, Mark J. Cameron, Alejandro B. Balazs, Stefan Gravenstein, Christopher L. King

## Abstract

The SARS-CoV-2 pandemic impact on nursing home (NH) residents prompted their prioritization for early vaccination. To fill the data gap for vaccine immunogenicity in NH residents, we examined antibody levels after BNT162b2 mRNA vaccine to spike, receptor binding domain (RBD) and for virus neutralization in 149 NH residents and 111 health care worker controls. SARS-CoV-2-naive NH residents mount antibody responses with nearly 4-fold lower median neutralization titers and half the anti-spike level compared to SARS-CoV-2-naive healthcare workers. By contrast, SARS-CoV-2-recovered vaccinated NH residents had neutralization, anti-spike and anti-RBD titers similar to SARS-CoV-2-recovered vaccinated healthcare workers. NH residents’ blunted antibody responses have important implications regarding the quality and durability of protection afforded by neoantigen vaccines. We urgently need better longitudinal evidence on vaccine effectiveness specific to NH resident populations to inform best practices for NH infection control measures, outbreak prevention and potential indication for a vaccine boost.

## Main text

The SARS-CoV-2 pandemic has severely affected nursing home (NH) residents prompting their prioritization for early vaccination. Recent reports show the BNT162b2 mRNA vaccine reduces COVID-19 hospitalization and mortality ^1,2 3,4^. However, no BNT162b2 mRNA vaccine phase 3 trial immunogenicity data exists for NH residents; their multiple morbidities, medications, and extreme age may blunt their immune response. Here, we examined antibody response to BNT162b2 mRNA vaccine in NH residents.

We obtained study approval from the New England IRB and consented subjects per protocol. We recruited NH residents for vaccination with and without prior SARS-CoV-2 infection (age 48-99, median age 76; n=149) and control comparators that included 1) healthcare workers (age 26-78; median age 48; n=111) with and without prior SARS-CoV-2 infection and, 2) an unvaccinated convalescent group 29-94 days after SARS-CoV-2 infection (age range 25-61, median age 53; n=22) with asymptomatic or mild disease (Table 1). All NH residents and 72 of 111 healthcare worker controls provided baseline blood samples 1-14 days before BNT162b2 mRNA vaccination and all were sampled 14±3 days after their second dose. We evaluated IgG to spike protein and its receptor binding domain (RBD) using bead-multiplex immunoassay and serum neutralization titers in a SARS-CoV-2-pseudovirus neutralization assay^5^.

**Table 1.** Demographics of Cohorts.

|  | **Control,**<br>**COVID**<br>**naïve**<br>**(n = 76)** | **Control,**<br>**prior**<br>**COVID**<br>**(n = 35)** | **NH,**<br>**COVID**<br>**naïve**<br>**(n = 90)** | **NH, prior**<br>**COVID**<br>**(n = 59)** | **Unvaccinated**<br>**comparator**<br>**(n = 22)** |
| --- | --- | --- | --- | --- | --- |
| Age: mean +/- SD | 49 +/- 11 | 48 +/- 12 | 78 +/- 11 | 78 +/- 10 | 46 +/- 13 |
| Age: Median (IQR) | 48 (39, 56) | 49 (38, 56) | 76 (70, 87) | 77 (71, 87) | 53 (36, 56) |
| Age: Range | 26-78 | 30-78 | 48-99 | 61-99 | 25-61 |
| Race & Ethnicity:<br>White,non-Hispanic | 62 (82%) | 29 (83%) | 79 (88%) | 50 (85%) | 17 (77%) |
| Race & Ethnicity:<br>African American,<br>non-Hispanic | 5 (7%) | 6 (17%) | 9 (10%) | 9 (15%) | 5 (23%) |
| Race & Ethnicity:<br>Hispanic | 4 (5%) | 0 (0%) | 1 (1%) | 0 (0%) | 0 (0%) |
| Race & Ethnicity:<br>Asian | 5 (7%) | 0 (0%) | 1 (1%) | 0 (0%) | 0 (0%) |
| Gender: Male | 39 (51%) | 13 (37%) | 57 (63%) | 35 (59%) | 8 (36%) |
| Gender: Female | 37 (49%) | 22 (63%) | 33 (37%) | 24 (41%) | 14 (64%) |

Figure 1 compares NH residents’ post-vaccination antibody with younger healthcare worker controls, stratified by prior SARS-CoV-2 infection as determined by antibody and/or PCR by the time of vaccination. SARS-CoV-2-naive NH residents mount antibody responses with nearly 4-fold lower median neutralization titers (521 vs 135, p<0.0001) and half the anti-spike level (4177 vs 1029, p<0.0001) compared to SARS-CoV-2-naive healthcare workers. After vaccination, 17% of SARS-CoV-2-naive NH residents have neutralizing titers at or below the lower limit of detection with only 1.3% of SARS-CoV-2-naive healthcare workers that low. Similarly, fewer vaccinated SARS-CoV-2-naive NH residents compared to vaccinated SARS-CoV-2 naive controls (36% vs 85%) have neutralization titers above the median of the unvaccinated SARS-CoV-2 convalescent younger adults. By contrast, neutralization, anti-spike and anti-RBD titers in SARS-CoV-2-recovered NH residents receiving vaccine were similar to SARS-CoV-2-recovered vaccinated healthcare workers (p>0.24 in all). NH residents’ blunted antibody responses have important implications regarding the quality and durability of protection afforded by neoantigen vaccines. We urgently need better longitudinal evidence on vaccine effectiveness specific to NH resident populations to inform best practices for NH infection control measures, outbreak prevention and potential indication for a vaccine boost.

**Figure 1.**
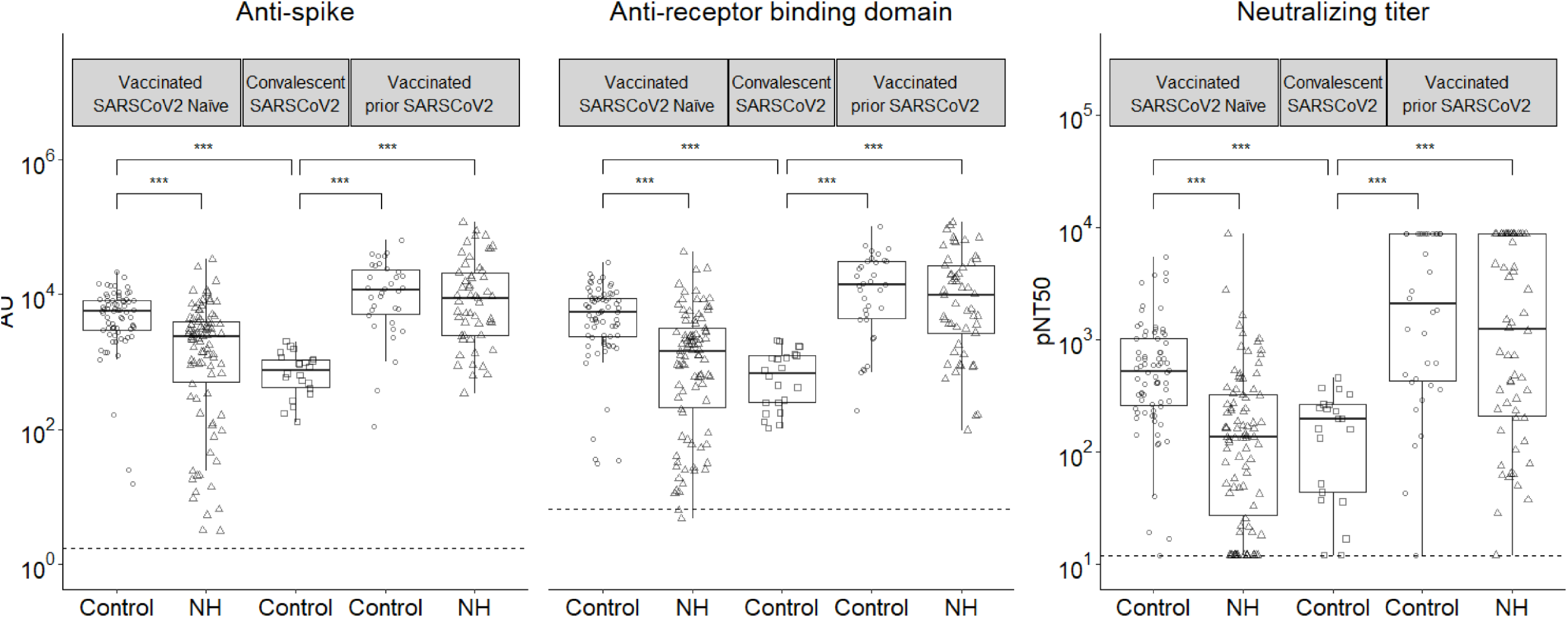
Humoral immune assessment of BNT162b2 mRNA vaccine vaccination in NH residents. Post-vaccination anti-spike, anti-RBD and serum neutralization titers are shown. On the x axis, NH refers to NH residents and Control refers to the vaccinated younger healthcare workers or unvaccinated SARS-CoV-2 convalescent individuals. The dotted line in each panel is the median pre-immunization value in the SARS-CoV-2-naive subjects. Anti-Spike and Anti-RBD differences in geometric means were assessed using t-tests of log-transformed values. Given observed points at upper and lower limits of detection in neutralizing titer assay (12-8748), differences in distribution were assessed using the Wilcoxon rank-sum test. P-values were adjusted within assay (n = 6 tests per assay, Bonferroni method), and adjusted p-values < 0.001 are indicated with stars (***). We did not detect significant differences in any assay between the NH and Control subjects with prior SARS-CoV-2 or between the SARS-CoV-2-naive NH subjects and the convalescent unvaccinated controls.

Support from NIH (AI129709-03S1, 3P01AG027296-11S1, and U01 CA260539-01) and VA

## Data Availability

Once the paper is in press we will release data as required by NIH with work that they fund.

